# The emergence and ongoing convergent evolution of the N501Y lineages coincides with a major global shift in the SARS-CoV-2 selective landscape

**DOI:** 10.1101/2021.02.23.21252268

**Authors:** Darren P Martin, Steven Weaver, Houryiah Tegally, Emmanuel James San, Stephen D Shank, Eduan Wilkinson, Alexander G Lucaci, Jennifer Giandhari, Sureshnee Naidoo, Yeshnee Pillay, Lavanya Singh, Richard J Lessells, NGS-SA, COVID-19 Genomics UK (COG-UK), Ravindra K Gupta, Joel O Wertheim, Anton Nekturenko, Ben Murrell, Gordon W Harkins, Philippe Lemey, Oscar A MacLean, David L Robertson, Tulio de Oliveira, Sergei L Kosakovsky Pond

**Author notes:** Full list of consortium names and affiliations are in the appendix (NGS-SA). Full list of consortium names and affiliations are in the appendix (COVID-19 Genomics UK (COG-UK)).

## Abstract

The emergence and rapid rise in prevalence of three independent SARS-CoV-2 “501Y lineages’’, B.1.1.7, B.1.351 and P.1, in the last three months of 2020 prompted renewed concerns about the evolutionary capacity of SARS-CoV-2 to adapt to both rising population immunity, and public health interventions such as vaccines and social distancing. Viruses giving rise to the different 501Y lineages have, presumably under intense natural selection following a shift in host environment, independently acquired multiple unique and convergent mutations. As a consequence, all have gained epidemiological and immunological properties that will likely complicate the control of COVID-19. Here, by examining patterns of mutations that arose in SARS-CoV-2 genomes during the pandemic we find evidence of a major change in the selective forces acting on various SARS-CoV-2 genes and gene segments (such as S, nsp2 and nsp6), that likely coincided with the emergence of the 501Y lineages. In addition to involving continuing sequence diversification, we find evidence that a significant portion of the ongoing adaptive evolution of the 501Y lineages also involves further convergence between the lineages. Our findings highlight the importance of monitoring how members of these known 501Y lineages, and others still undiscovered, are convergently evolving similar strategies to ensure their persistence in the face of mounting infection and vaccine induced host immune recognition.

---

[BODY]

## Introduction

In the first eleven months of the SARS-CoV-2 pandemic (December 2019 - October 2020), the evolution of the virus worldwide was in the context of a highly susceptible new host population (Dearlove et al., 2020; MacLean et al., 2021). Other than the early identification of the D614G substitution in the viral spike protein (Korber et al., 2020; Plante et al., 2021; Zhang et al., 2020) and P323L in the viral RNA dependent RNA polymerase protein (Garvin et al., 2020), both of which may have increased viral transmissibility without impacting pathogenesis (reviewed in (Peacock et al., 2021)), few mutations were epidemiologically significant and the evolutionary dynamics of the virus were predominantly characterized by a mutational pattern of slow and selectively-neutral random genetic drift (Dearlove et al., 2020; MacLean et al., 2021). This behavior is consistent with exponential growth in a population of naive susceptible hosts that do not exert significant selective pressures on the pathogen prior to transmission events (MacLean et al., 2021). Past pandemics and long-term evolutionary dynamics of RNA viruses attest to the fact that such an evolutionary “lull” does not necessarily last. Indeed, in late 2020, three relatively divergent SARS-CoV-2 lineages emerged in rapid succession: (i) alpha, B.1.1.7 or 501Y.V1 which will hereafter be referred to as V1 (Rambaut et al., 2020a) (ii) beta, or 501Y.V2 which will hereafter be referred to as V2 (Tegally et al., 2021) and (iii) gamma, P.1 or 501Y.V3 which will hereafter be referred to as V3 (Faria et al., 2021).

Viruses in each of the three lineages (which will hereafter be collectively referred to as 501Y lineages) have multiple signature (or lineage defining) deletions and amino acid changing substitutions (Figure 1), many of which impact key domains of the spike protein: the primary target of both infection and vaccine induced immune responses. Prior to this, while many distinct spike mutations had been observed, all circulating SARS-CoV-2 lineages were defined by small numbers of mutations. All of the 501Y lineages also have significantly altered phenotypes: increased human ACE2 receptor affinity (V1, V2 and V3) (Nelson et al., 2021; Starr et al., 2020; Zahradnik et al., 2021), increased transmissibility (V1, V2 and V3) (Althaus et al., 2021; Faria et al., 2021; Lubinski et al., 2021; Pearson et al., 2021; Public Health England, 2020; Volz et al., 2021), substantially increased capacity to overcome prior infection and/or vaccination-induced immunity (V2 and V3) (Cele et al., 2021; Garcia-Beltran et al., 2021; Hoffmann et al., 2021; Shinde et al., 2021; Wibmer et al.; Wu et al., 2021) and associations with increased virulence (V1 and V3) (Faria et al., 2021; Horby et al., 2021). Why did the heavily mutated 501Y lineages all arise on different continents at almost the same time? Was it due to an intrinsic change in the capacity of SARS-CoV-2 to adapt, or was it a shift in the host selective environment extrinsic to the virus?

**Figure 1.**
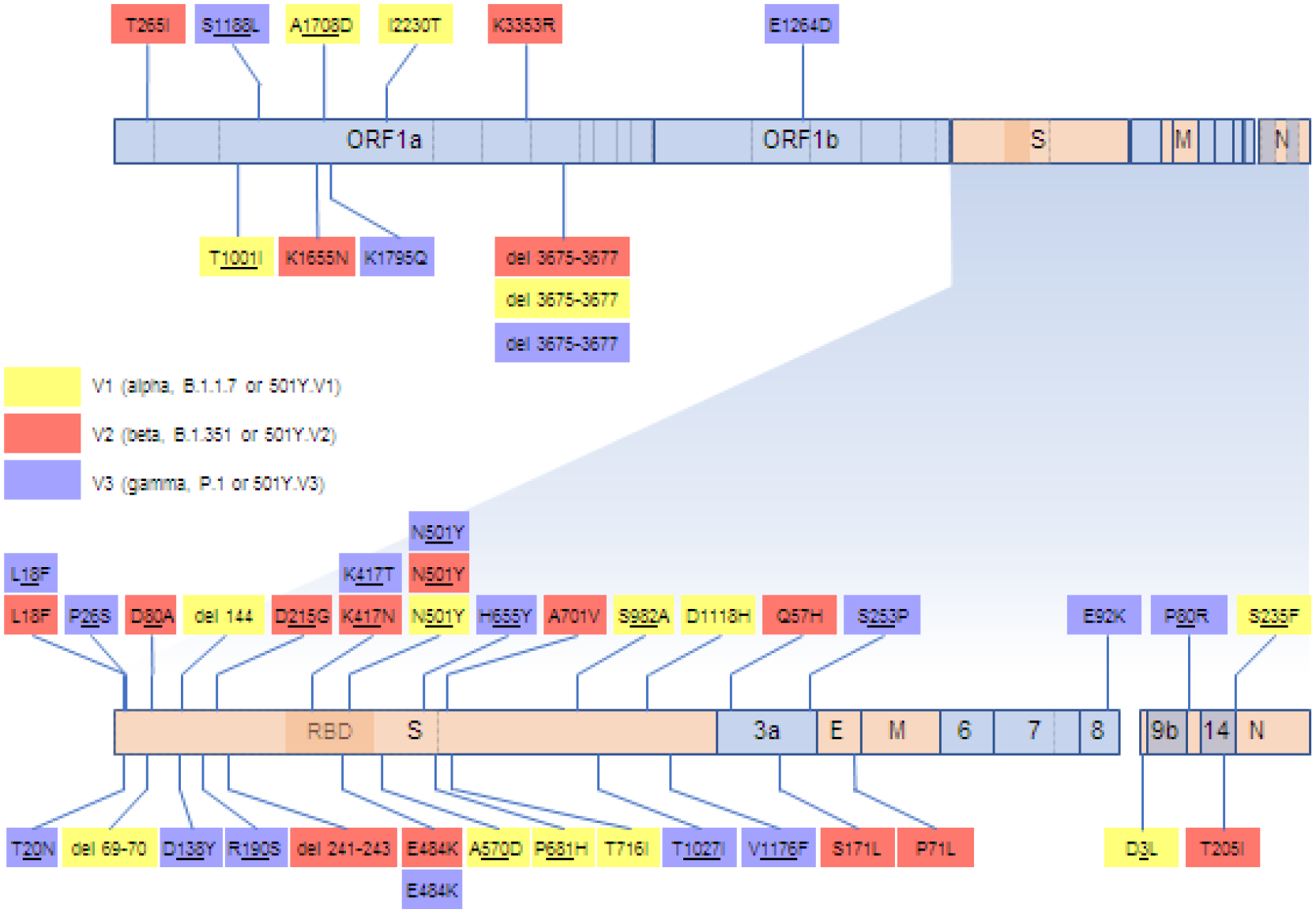
SARS-CoV-2 genome map indicating the locations and encoded amino acid changes of what we considered here to be signature mutations of V1, V2 and V3 sequences. Genes represented with light blue blocks encode non-structural proteins and genes in orange encode structural proteins:S encodes the spike protein, E the envelope protein, M the matrix protein, and N the nucleocapsid protein. Within the S-gene, the receptor binding domain (RBD) is indicated by a darker shade and the site where the S protein is cleaved into two subunits during priming for receptor binding and cell entry is indicated by a dotted vertical line.

Evidence that natural selection has played a pivotal role in the emergence of V1, V2 and V3 can be found in the remarkable patterns of independently evolved convergent mutations that have arisen within the members of these lineages (Figure 1; (Peacock et al., 2021)). One of the most striking of these parallel changes is a nine nucleotide deletion between genome coordinates 11288 and 11296 (here and hereafter all nucleotide and amino acid coordinates refer to the GenBank reference genome NC_045512). This deletion is within the portion of ORF1ab that encodes non-structural protein 6 (nsp6): a component of the SARS-CoV-2 membrane-tethered replication complex that likely influences the formation and maturation of autophagosomes (Cottam et al., 2011), and decreases the effectiveness of host innate antiviral defences by reducing the responsiveness of infected cells to, and antagonizing the production of, type I interferons (Lei et al., 2020; Miorin et al., 2020; Xia et al., 2020). Relative to some of the earliest characterized SARS-CoV-2 A and B variants, V1 and V2 viruses have demonstrably less sensitivity to type I interferons (Guo et al., 2021; Thorne et al., 2021) and V1 displays greater antagonism of type I interferon mediated immune activation during the early stages of infection in cultured lung epithelial cells (Thorne et al., 2021). However, it remains unknown whether these characteristics of V1 and V2 viruses are in any way attributable to their shared 11288-11296 deletion. Independently evolved instances of this deletion have been repeatedly found prior to the emergence of the 501Y lineages and identical independently evolved deletions are also found together with other 501Y lineage signature mutations in the SARS-CoV-2 lineages, B.1.620 (Dudas et al., 2021), B.1.1.318, B.1.525 (https://github.com/cov-lineages/pango-designation/issues/4) and B.1.526 (Annavajhala et al., 2021). This degree of convergent evolution implies that, in the context of the B.1.620, B.1.1.318, B.1.525, B.1.526 and the 501Y lineages at least, the 11288-11296 deletion is likely highly adaptive.

Additionally, there are four convergent spike gene mutations that are each shared between members of different 501Y lineages. Almost all the spike genes of sequences in these lineages carry the N501Y mutation at a key receptor binding domain (RBD) site that increases the affinity of the spike protein for human ACE2 receptors by ∼2.1-3.5 fold (Starr et al., 2020; Yuan et al., 2021; Zahradnik et al., 2021). The vast majority of V2 and V3 variants and ∼0.3% of more recent samples of V1 variants also have a spike E484K mutation. Whereas in the presence of 501N, 484K has a modest positive impact on ACE2 binding (Starr et al., 2020), when present with 501Y, these mutations together synergistically increase ACE2-RBD binding affinity ∼12.7 fold (Nelson et al., 2021; Zahradnik et al., 2021). Crucially, E484K and other mutations at S/484 also frequently confer protection from neutralization by both convalescent sera (Greaney et al., 2021a), vaccine elicited antibodies (Collier et al., 2021; Wang et al., 2021a, 2021b; Wu et al., 2021), and some monoclonal antibodies (Greaney et al., 2021a; Starr et al., 2021; Wang et al., 2021b). There is therefore increasing evidence that viruses carrying the E484K mutation (with or without 501Y) will be able to more frequently infect both previously infected and vaccinated individuals (Collier et al., 2021; Wang et al., 2021a, 2021b; Wu et al., 2021).

A third RBD site that is mutated in both V2 and V3 is S/417. Whereas V2 sequences generally carry a K417N mutation, V3 sequences carry a K417T mutation. Although both the K417N and K417T mutations can reduce the affinity of spike for ACE2, in conjunction with the N501Y and E484K mutations ACE2 binding is restored to that of wild-type Spike (Yuan et al., 2021). K417N and K417T also both have moderately positive impacts on spike expression (Starr et al., 2020) and these and other mutations at S/417 provide modest protection from neutralization by some convalescent sera (Greaney et al., 2021a; Wang et al., 2021b), vaccine induced antibodies (Wang et al., 2021b) and some neutralizing monoclonal antibodies (Starr et al., 2021; Wang et al., 2021b; Wibmer et al.).

A fourth spike gene mutation that is shared by ∼48% of V2 sequences and by all V3 sequences is L18F. This amino acid change is predicted to have a modest impact on the structure of spike (Nguyen et al., 2021) and also protects from some neutralizing monoclonal antibodies (McCallum et al., 2021). Viruses carrying the L18F mutation increased in prevalence from the start of the pandemic and now account for ∼10% of sampled SARS-CoV-2 sequences.

These five convergent mutations in different rapidly spreading SARS-CoV-2 lineages is compelling evidence that they each, either alone or in combination, provide some significant fitness advantage. The individual and collective fitness impacts of the other signature mutations in V1, V2 and V3 remain unclear. A key way to infer the fitness impacts of these mutations is to examine patterns of synonymous and non-synonymous substitutions at the codon-sites where the mutations occurred (Kosakovsky Pond and Frost, 2005). Specifically, it is expected that the most biologically important of these mutations will have occurred at codon-sites that display substitution patterns across the wider SARS-CoV-2 phylogeny that are dominated by non-synonymous mutations (i.e. mutations that alter encoded amino acid sequences); patterns that are indicative of positive selection.

Here, using a suite of phylogenetics-based natural selection analysis techniques, we examine patterns of positive selection within the protein coding sequences of viruses in the V1, V2 and V3 lineages which, together with mutation frequency changes over time, we use to identify the specific mutations that are at present most likely contributing to the increased adaptation of these lineages. We find that the emergence of the 501Y lineages coincided with a marked global change in positive selection signals, indicative of a general shift in the selective environment within which SARS-CoV-2 is evolving. Against this backdrop, the 501Y lineages all display evidence of substantial ongoing adaptation involving in many cases mutations at positively selected genome sites that both converge on mutations seen in other 501Y lineages, and are rapidly rising in frequency in different lineages. This pattern suggests that viruses in all three lineages are presently still climbing very similar adaptive peaks, and, therefore, that viruses in all three lineages are likely in the process of converging on a similar adaptive endpoint.

## Results and Discussion

### There has been a recent detectable shift in selective pressures acting on circulating SARS-CoV-2 variants

Analyses of positive selection on SARS-CoV-2 genomes undertaken prior to the emergence of 501Y lineages revealed mutational patterns dominated by neutral evolution (MacLean et al., 2021). There were, however, indications that some sites in the genome had experienced episodes of positive selection (Garvin et al., 2020; Korber et al., 2020; Plante et al., 2021; Zhang et al., 2020). Through regular analyses of global GISAID data (Elbe and Buckland-Merrett, 2017) starting in March 2020, we tracked the extent and location of positive and negative selective pressures on SARS-CoV-2 genomes (Figure 2). The power of these analyses to detect evidence of selection acting on individual codon-sites progressively increased over time with rising numbers of sampled genome sequences and sequence diversification.

**Figure 2.**
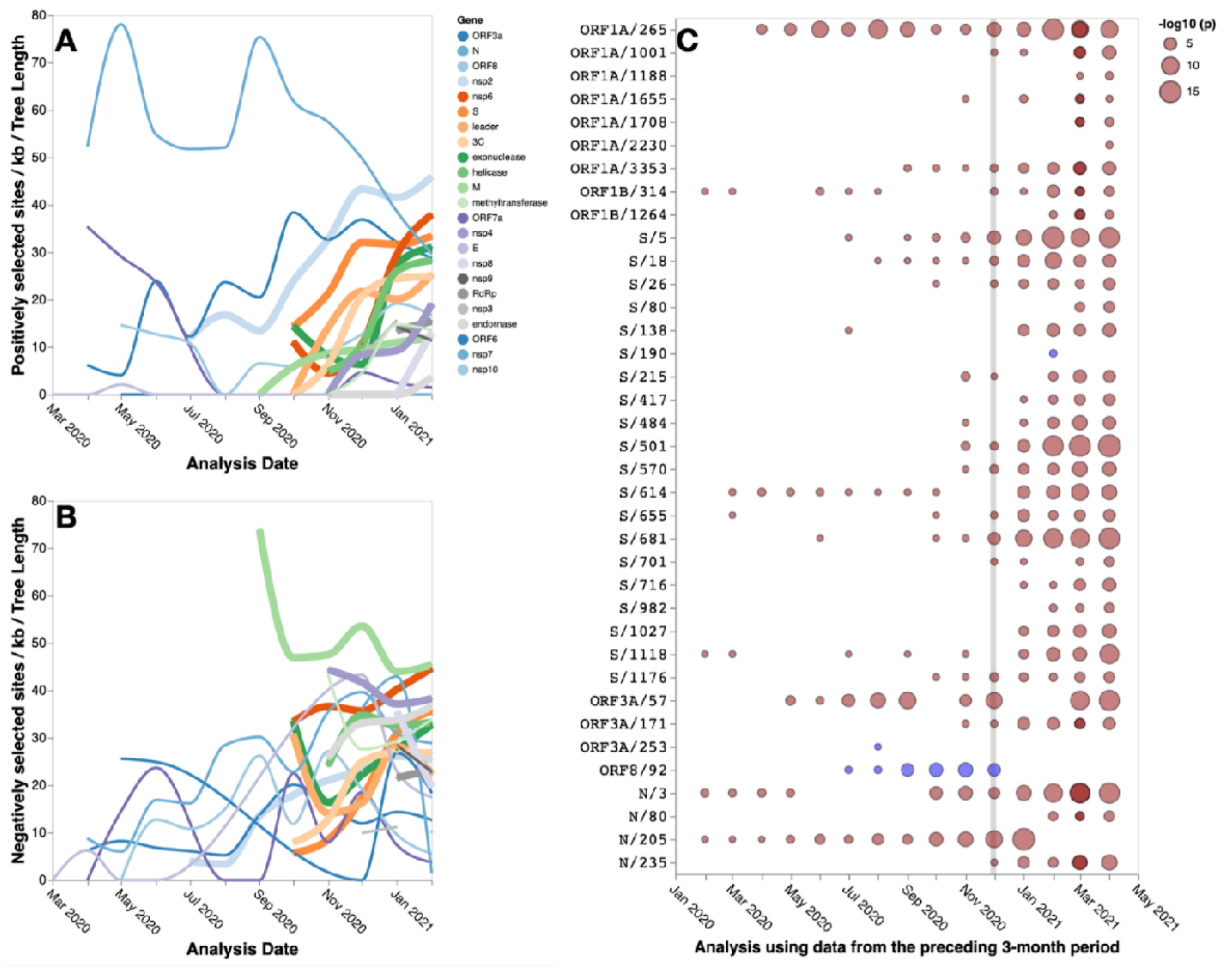
Signals of positive and negative selection at individual codon-sites that were detectable with the FEL method at different times between March 2020 and April 2021, applied to sequences sampled over 90-day intervals; the plotted date shows the end of the 90-day period. **A/B**. The gene-by-gene/per Kb/per unit tree length density of codons detectably evolving under positive/negative selection between March 2020 and February 2021. Whereas genes for which the maximum observed density of positively/negatively selected sites was reached in February 2021 are shown with thicker lines, genes with associated trees that have a total length shorter than 0.5 subs/site for a given time-period are not shown. A version of panel A with all genes displayed separately is given in Figure S1. **C** Signals of positive selection detected at 37 V1, V2 and V3 signature mutation sites between March 2020 and April 2021. Also included for reference are sites previously detected to be evolving under positive selection such as S/614, the site of the D614G mutation that is present in all three of the 501Y lineages, S/5 and RdRp/P323L (ORF1b/314). Circles indicate the statistical significance of the FEL test with red indicating positive selection and blue indicating negative selection. The vertical line indicates December 1st, 2020; the approximate date when the importance of the V1 and V2 lineages was first noticed.

Even accounting for this expected increased power of detection, it is evident that a significant shift in selective pressures occurred ∼11 months after SARS-CoV-2 cases were first reported in Wuhan City in December 2019. Specifically, during November 2020 this change in selection pressures manifested in substantial increases in the numbers of SARS-CoV-2 codon-sites that were detectably evolving under both positive and negative selection. This increase accelerated through February 2021, with sites found to be evolving under diversifying positive selection in several genomic regions (p ≤ 0.01 with the FEL selection detection method (Kosakovsky Pond and Frost, 2005)) rapidly increasing in density for several key genes including S, nsp2, and nsp6 (Figure 2A; Figure S1). This increase cannot be fully explained by increased sampling at later time points, as our density measurements correct for the increased phylogenetic signal, measured in the total length of internal tree branches (MacLean et al., 2020). This sudden increase in the density of sites that were detectably evolving under positive selection coincided with epidemic surges in multiple parts of the world in both hemispheres, many of which were driven by the emerging V1, V2 and V3 lineages.

Among the 37 signature mutation sites in V1, V2 and V3 (Figure 1), 14 were detectably evolving under positive selection in November 2020 whereas this number increased to 22 by January 2021, and 30 by April 2021 (Figure 2C). The only signature mutation shared between any of the three 501Y lineages that was detectably evolving under positive selection before November 2020 was S/18 which was detected for the first time in August 2020.

Our regular tracking of positively selected SARS-CoV-2 codon-sites prior to November 2020 therefore yielded no clear indications that non-synonymous substitutions at the crucial RBD sites S/417, S/484 or S/501 (the other key convergent signature mutation sites in the 501Y lineages), provided SARS-CoV-2 with any substantial fitness advantages in the first 11 months of the pandemic. Instead, the sporadic weak selection signals that these analyses yielded between July and November were of adaptive amino acid substitutions in the Spike N-terminal domain (S/18, and the V3 signature sites, S/26 and S/138), near the furin cleavage site (the V3 signature site, S/655 and the V1 signature site S/681), and in the C-terminal domain (the V1 signature site, S/1118 and the V3 signature site S/1176). Conversely, for much of the latter half of 2020 the relatively strong and consistently detected selection signals at the V1 signature site, N/3, and the V2 signature sites, ORF1ab/265 (nsp2 codon 85) and ORF3a/57, clearly indicated that some substitutions at these sites were likely at least mildly adaptative.

Taken together, these patterns of detectable selection suggest that the adaptive value of signature 501Y lineage RBD mutations may have only manifested after a selective shift that occurred shortly before November 2020.

### Signals of selection within the V1, V2 and V3 lineages up until March 2021

We initially restricted our selection analyses to only consider mutations arising within V1, V2 and V3 lineage sequences sampled before April 2021 to assess the adaptive processes at play within these lineages during and immediately after the perceived shift in the SARS-CoV-2 selective landscape in late 2020. We were specifically interested in identifying positive selection signals at individual codon-sites that reflected these “early” selective processes.

We collected all sequences assigned to B.1.1.7 (V1), B.1.351 (V2), and P1 (V3) PANGO lineages (Rambaut et al., 2020b) in GISAID (Elbe and Buckland-Merrett, 2017) as of 20 April 2021 and tested these sequences for evidence of positive selection at individual codon-sites using MEME (Murrell et al., 2012) and FEL (Pond et al., 2006). Both of these methods were restricted to analysing mutations that mapped to internal branches of V1, V2 and V3 phylogenetic trees: i.e. mutations that almost certainly would have only arisen before mid-March 2021.

These analyses revealed evidence of positive selection at 151 individual codon-sites (at p < 0.05) across all lineages including 80 in V1, 41 in V2, and 37 in V3 (Table S1). This is indicative of substantial adaptation of V1, V2 and V3 sequences between the time of their emergence and March 2021.

### Signals of ongoing mutational convergence at signature mutation sites

Notable among the lineage-specific positive selection signals were 22/151 at lineage defining mutation sites: 8/11 of the V1 signature sites, 4/14 of the V2 ones and 13/17 of the V3 ones (Figure 3 and see underlined codon-site numbers in Figure 1). Given that (i) each lineage was defined by the signature mutations along the phylogenetic tree branch basal to its clade, and (ii) that these basal branches were included in the lineage-specific selection analyses, these selection analysis results were biased in favour of detecting the signature mutations as evolving under positive selection. We therefore used the selection results for signature mutation sites only to identify the signature mutations that, relative to the background reference sequences, were evolving under the strongest degrees of positive selection during or before March 2021.

**Figure 3.**
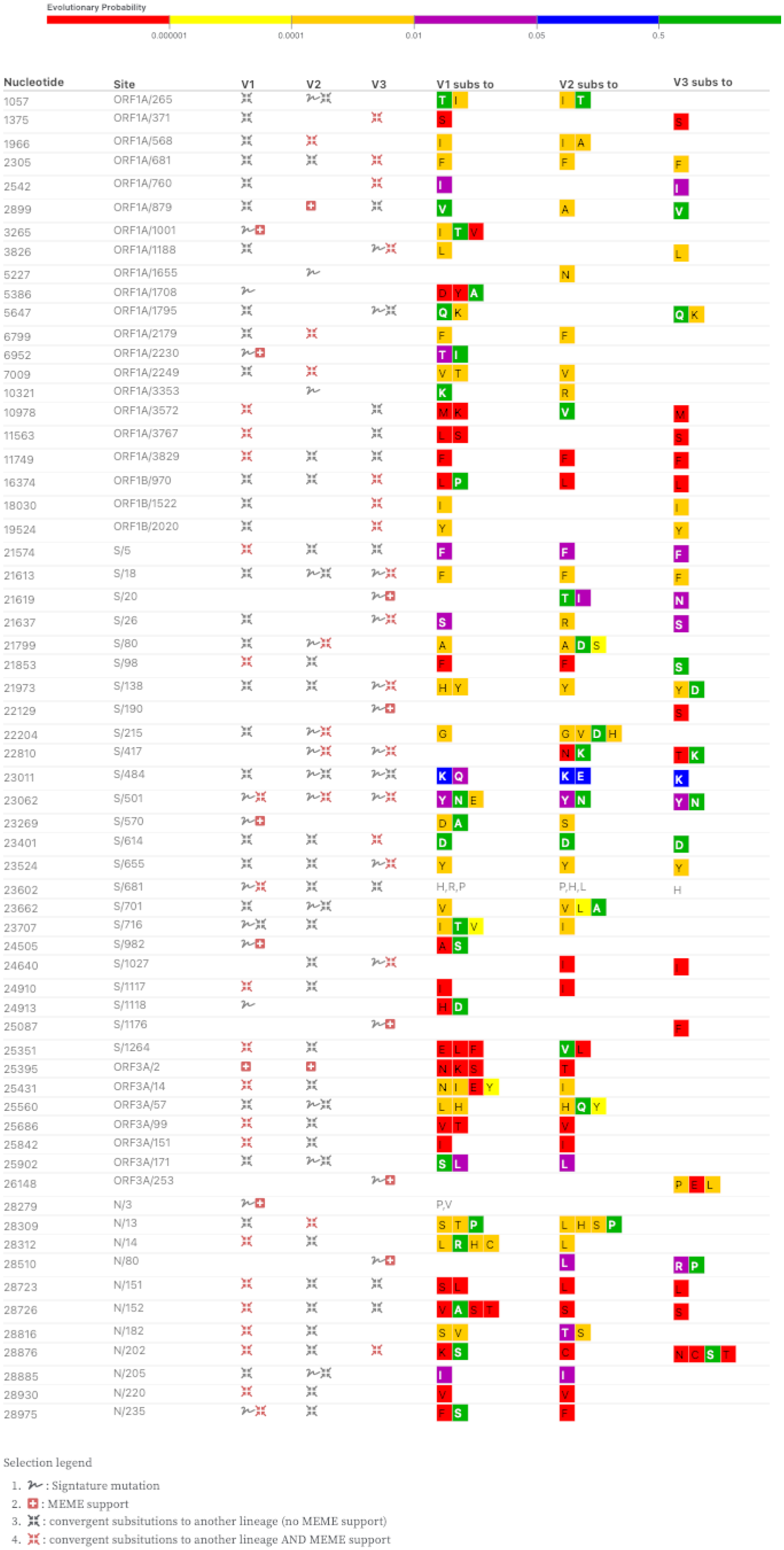
Genome sites where signature and convergent mutations occur within the 501Y lineage sequences. Sites detectably evolving under positive selection along internal branches (MEME p-value ≤ 0.05) of the V1, V2 and V3 phylogenies are indicated with red icons. We restricted our analysis ≤ to data collected up to April 2021 to focus on interpreting predictive positive selection signals arising from mutations occurring before March 2021, which could then be corroborated by examining mutation frequency data from later months. Labels within the coloured blocks indicate amino acid substitutions with block colours indicating model-based predictions of the probable evolutionary viability of the observed amino acid substitutions based on the numbers of times these substitutions have been observed in related coronaviruses that infect other host species. The absence of colour indicates unprecedented substitutions, red indicates highly unusual substitutions and green indicates common substitutions seen at homologous sites in non-SARS-CoV-2 coronaviruses. ORF8 signals have been excluded.

Most noteworthy of the 22 signature mutation sites that displayed the strongest evidence of lineage-specific positive selection are codons S/18, S/80, S/417, S/501, S/655 and S/681 in that all are either suspected or known to harbour mutations with potentially significant fitness impacts (Garry et al., 2021; Greaney et al., 2021b, 2021a; Lubinski et al., 2021; McCallum et al., 2021; Starr et al., 2020; Wang et al., 2021a, 2021b; Zahradnik et al., 2021).

Before March 2021 there were particularly interesting mutational dynamics at codon S/18 in the V1 lineage. Whereas the L18F mutation is almost fixed in all currently sampled V3 lineage sequences, it occurred (and persisted in descendent variants) at least twice in the V1 lineage and at least four times in the V2 lineage. S/18 falls within multiple different predicted CTL epitopes (Campbell et al., 2020) and the L18F mutation is known to reduce viral sensitivity to some neutralizing monoclonal antibodies (McCallum et al., 2021). An F at residue S/18 is also observed in 10% of other known Sarbecoviruses and the L18F mutation was the 28th most common in sampled SARS-CoV-2 genomes on 04 June 2021. Having occurred independently numerous times since the start of the pandemic, S/18 has also been detectably evolving under positive selection in the global SARS-CoV-2 genome dataset since August 2020 (Figure 2C).

Similar convergence patterns to those observed at S/18 could be seen at 17 other signature mutation sites that, before March 2021, were detectably evolving under positive selection in either the global (Figure 2C) or lineage-specific (Figure 3) datasets: ORF1a/265, ORF1a/1188, S/26, S/138, S/215, S/417, S/484, S/501, S/655, S/681, S/701, S/716, S/1027, S/1176, ORF3a/57, N/205, and N/235.

Of these, S/655, S/681, S/701 and S/716 are noteworthy in that they fall within 30 residues of the biologically important Spike protein furin cleavage site (S/680 to S/689). Whereas some V2 and V3 sequences had, by March 2021, independently acquired the signature V1 mutations, P681H and T716I, some V1 and V2 sequences had independently acquired the V3 signature mutation, H655Y, and some V1 sequences had independently acquired the V2 signature mutation, A701V. Additionally, whereas a convergent A701V mutation is also found in the B.1.526 and S/E484K carrying lineage that was first identified in New York (Annavajhala et al., 2021), P681H is found in the S/E484K and S/N501Y carrying P.3 lineage first identified in the Philippines (Tablizo et al., 2021), and both S/H655Y and S/P681H are found in the highly mutated S/E484K carrying A.VOI.V2 lineage first identified in Tanzanian travellers (de Oliveira et al., 2021).

Any of H655Y, P681H, A701V or T716I might directly impact the efficiency of viral entry into host cells (Garry et al., 2021). SARS-CoV-2 variants with deletions of the furin cleavage site have reduced pathogenicity (Johnson et al., 2021; Lau et al., 2020) and the P681H mutation - which falls within this site - likely increases the efficiency of furin cleavage by replacing a less favourable uncharged amino acid with a more favourable positively charged basic one (Garry et al., 2021; Lubinski et al., 2021). Whereas sites S/655, and S/681 are also detectably evolving under positive selection in at least one of the lineage specific datasets, S/655, S/681, A/701 and S/716 are all detectably evolving under positive selection in the March and April 2021 global SARS-CoV-2 datasets; important additional indicators that are consistent with the H655Y, P681H, A701V and T716I mutations being adaptive.

### Non-convergent mutations at signature mutation sites might still be evolutionarily convergent

In addition to the 17 signature mutation sites displaying evidence of both convergent mutations between the V1, V2 and V3 lineages, and positive selection in the global and/or lineage-specific datasets, four signature mutation sites with lineage-specific signals of positive selection (S/20, S/138, S/215 and S/570; Figure 1) display evidence of predominantly divergent mutations (at the amino acid replacement level), where the same site is mutated as in another lineage, but to a different amino acid (Figure 3). A fifth site, S/80, displays evidence of both convergent and divergent mutations. All but S/20 are also detectably evolving under positive selection in the global SARS-CoV-2 dataset (Figure 2).

The diverging mutations at these five sites might in fact also be contributing to the overall patterns of evolutionary convergence between the lineages; just via different routes. Four of these sites (S/20, S/80, S/138, and S/215) fall within a portion of the spike N-terminal domain that is an “antigenic supersite” targeted by multiple monoclonal and infection-induced neutralizing antibodies (McCallum et al., 2021). It is therefore plausible that these sites are evolving under immunity-driven diversifying selective pressures. In this regard, while mutations at S/20, S/80, S/138, S/215 and S/570 in different lineages do not predominantly converge on the same encoded amino acid states, they could nevertheless still be convergent on similar fitness objectives (immune escape or compensation for the fitness costs of other mutations): such as is likely the case with the also not-strictly-convergent V2 K417N and V3 K417T signature mutations (Greaney et al., 2021b, 2021a; Nelson et al., 2021).

### Positive selection may be driving further convergence at non-signature mutation sites

In addition to lineage-specific signals of positive selection being detected at 22 of the signature mutation sites that characterize each of V1, V2 and V3 (Figure 1), such signals were also detected in lineage-specific datasets at 129 non-signature mutation sites (Table S1). As with the positively selected signature mutation sites, these selection signals are based on mutations that map to internal V1, V2 and V3 tree branches and likely reflect selective processes operating before mid-March 2021.

To test whether positive selection acting at these 129 codon-sites might have favoured convergent amino acid changes across the three lineages, we examined mutations occurring at these sites for evidence of convergence between two or more of the lineages. This revealed the occurrence of convergent mutations between sequences in different lineages at 28/129 (21.7%) of these sites, including ten in ORF1a, seven in the N-gene, five in the S-gene (Figure 3), three in ORF3A. and three in ORF1B (Figure 3).

The lineage-specific positive selection signals detected at these 28 sites reflect repeated convergent non-synonymous mutations within each lineage that likely increase the fitness of the genomes in which they occur. Accordingly, 18/28 of the codons where these inter- and intra-lineage convergent mutations occur were also detectably evolving under either pervasive or episodic positive selection within the April global SARS-CoV-2 dataset (IFEL p-values < 0.05; Figure S2). This concordance between the lineage-specific and global selection signals is strong evidence that an appreciable proportion of the convergent non-signature site mutations are broadly adaptive (as opposed to being only epistatically adaptive in the context of 501Y lineage virus genomes).

The degree of overlap between non-signature mutation sites detectably evolving under selection in the V1, V2, and V3 lineages and displaying evidence of inter-lineage convergent mutations (Figure 3 and Table S1) cannot be adequately explained by chance alone. Restricting ourselves only to variable sites that are shared between lineages (i.e. just those sites where it was possible to detect selection and/or convergent mutations) and the numbers of selected sites in each lineage, we conducted a permutation test for the numbers of detectable convergent, positively selected non-signature site mutations between each pair of lineages. Overlap by chance can be rejected for V1/V2 (p<0.001), V1/V3 (p=0.002), and V2/V3 (p=0.002). This pattern supports the hypothesis that all three lineages are at present accumulating convergent non-signature site mutations that are contributing to their ascent of the same fitness peak.

### Changes in mutation frequencies since mid-March 2021 corroborate the inferred fitness advantages of convergent mutations at positively selected genome sites

Although it is clear that in the regions of the world where the prevalence of 501Y lineage viruses have increased since December 2020, these viruses have had substantial fitness advantages over the SARS-CoV-2 variants that preceded them, it remains unclear what the precise biological advantages were. The two most likely, non-exclusive, reasons for their increased fitness are: (1) that they were better at infecting people that had been previously infected (V2 and V3) (Cele et al., 2021; Garcia-Beltran et al., 2021; Wibmer et al.; Wu et al., 2021) and/or (2) that they were more transmissible (V1, V2 and V3)(Faria et al., 2021; Pearson et al., 2021; Volz et al., 2021).

While it is likely that all the convergent mutations that are detectable at positively selected sites (Figure 3) impact SARS-CoV-2 transmissibility and/or immune escape in at least some specific situations, it remains unclear what the relative fitness impacts of particular mutations at these sites are. It is, however, expected that population-wide frequencies of newly arising mutations which directly contribute to increased fitness should, at least initially, increase at a rate which is proportional to the magnitude of their fitness contribution. We therefore tested for changes in the frequencies of inter-lineage convergent mutations at the 28 non-signature mutation sites represented in Figure 3, and the full complement of 32 signature substitution mutation sites found in the 501Y lineage viruses (Figure 1). Specifically, this involved partitioning V1, V2 and V3 sequences deposited in GISAID by 01 June into “early” and “late” datasets that respectively contained sequences sampled before and after 15 March 2021: a date by which all of the internal branch mutations that yielded the detectable positive selection signals in our lineage-specific selection analysis datasets (i.e. those represented in Figure 3 and Table S1) would have already arisen. The frequencies of mutations evident at the 32 signature and 28 non-signature mutation sites were then compared between each of the V1, V2 and V3 early and late dataset pairs.

Over twofold increases in frequency between 15 March and 01 June 2021 were detected in at least one of the three 501Y lineages for at least one of the observed mutations at 28/60 of the analysed genome sites (these increases are all statistically significant in 2×2 contingency tables using the conservative Bonferroni multiple testing correction). Among these 28 sites are 19 where mutations in one (at 14 sites) or two (at five sites) of the 501Y lineages first converged on a signature mutation that characterizes a different 501Y lineage, and then proceeded to double in frequency between 15 March and 01 June 2021 (Table S2): an indication that the convergence mutations at these 19 sites may have each provided a fitness advantage. Based on the observed degree of frequency increases, mutations such as ORF1a/1708D (corresponding to nsp3/890D with 15.8 and >12.0 fold increases in V2 and V3 respectively), S/26S (>13 fold increase in V2), S/716I (3.7 and >13.5 fold increases in V2 and V3 respectively), S/1027I (>44 fold increase in V2), S/1118H (4.0 and >20 fold increases in V2 and V3 respectively), S/1176F (19.5 fold increase in V2) and ORF3/171L (11.9 fold increase in V3) are the signature mutations that, in addition to the ORF1a/3675-3677Del, S/18F, S/417N/T E484K and S/501Y mutations, are likely to have the greatest positive impact on the fitness of the 501Y lineage viruses within which they occur.

Similarly, among the nine positively selected sites where non-signature mutations both converge between viruses in two or more of the 501Y lineages, and then more than double in frequency between 15 March and 01 June 2021, ORF1b/1522I (corresponding to Helicase/590I with 1.9, 12.7 and 4.8 fold increases in V1, V2 and V3 respectively), S/98F (2.5, 5.3 and >6.0 fold increases in V1, V2 and V3 respectively), and E71T/R (respectively 5.8 and >10 fold increases in V1) are likely the most fitness-enhancing mutations.

In total 19/47 of the analysed convergent mutations that were suggested by our global and lineage-specific positive selection analyses to have contributed to the fitness of 501Y lineage viruses prior to March 2021, more than doubled in frequency in at least one of the lineages between 15 March and 01 June 2021. The 01 June dataset revealed a further nine previously undetected convergent mutations at 501Y lineage signature sites (ORF1a/1001I, ORF1a/1655N, ORF1a/1708D, ORF1b/970, S/215, S/1118, S/1176F, E/71L and N/205; Table S2) that were associated with positive selection signals in the global dataset (Figure 4) and which also more than doubled in frequency between 15 March and June 2021. Of all the 501Y lineage mutations that have so far been observed, the convergent mutations at these 28 sites have the strongest corroborating evidence supporting their individual and/or collective contributions to the ongoing adaptation of 501Y lineage viruses during the present phase of the pandemic.

**Figure 4.**
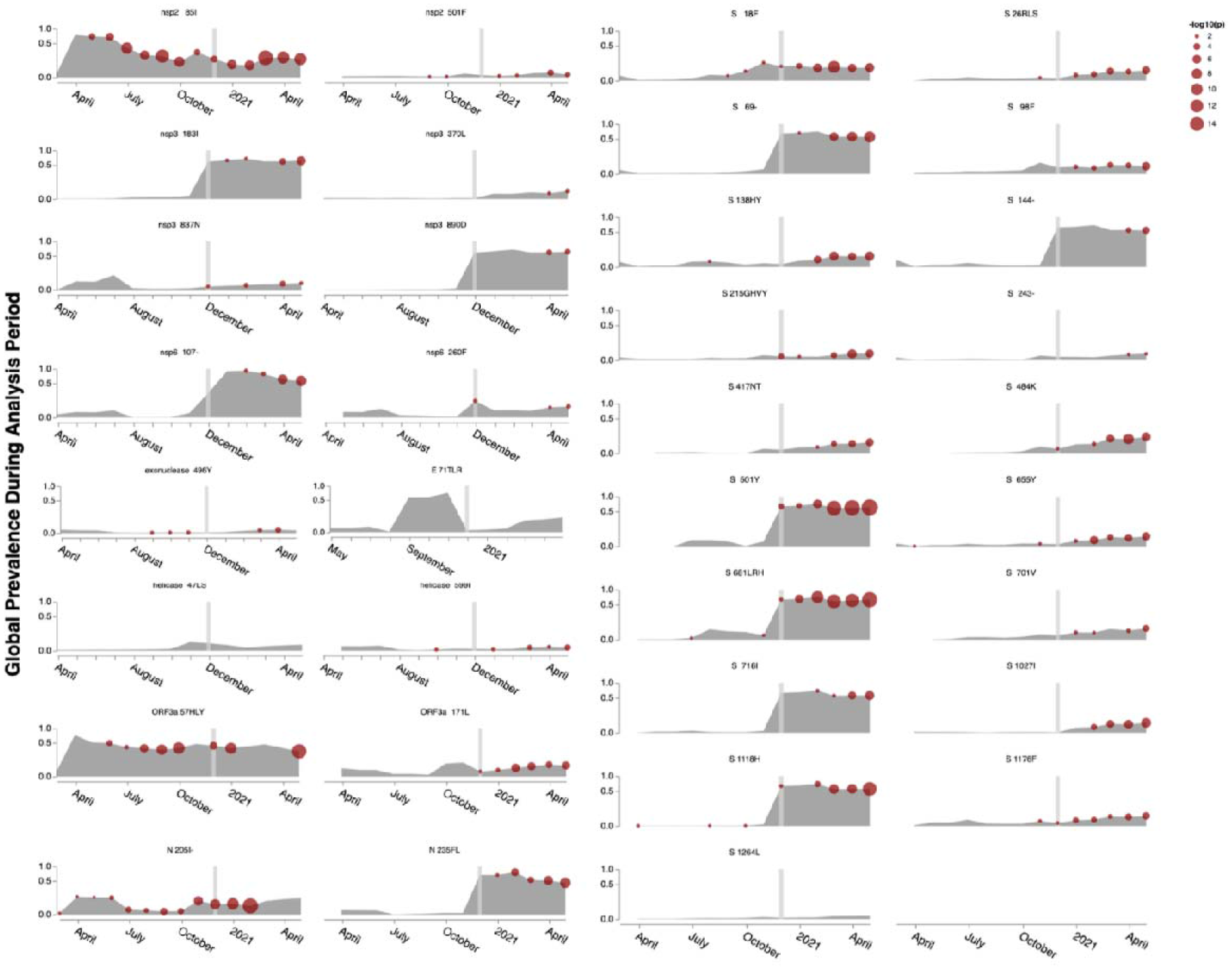
Selection signals evident in the global data at the subset of sites identified by the positive selection, convergence and mutation frequency change analyses as likely contributing to the fitness of the 501Y lineage viruses: a subset of sites and their associated amino acid states that we refer to as the 501Y lineage meta-signature. The strengths of detected selection signals (with the FEL method) are indicated by the sizes of the red dots. Selection tests were performed on sequence data collected within the preceding three months (i.e. red spots plotted in April reflect the analysis of sequences sampled between 01 January and 01 April). The vertical bar indicates 01 December 2020. The global frequencies of the represented mutations are indicated in grey. These frequencies are strongly biased by, and therefore track in many instances, the rapid rise of V1 viruses in the UK, Europe and North America: the regions of the world responsible for >90% of all SARS-CoV-2 genome sequencing since January 2001.

### Where is the evolution of the 501Y lineages headed?

Regardless of how exactly each of these 28 convergent mutations impact the fitness of 501Y lineage viruses, it is apparent that the evolution of these viruses will likely involve further selection-driven mutational convergence at these sites both between viruses within individual lineages, and between viruses in the different lineages. Based on our selection analyses, the convergence patterns that we have so-far detected, and the rises in frequencies between 15 March and 01 June 2021 of 501Y viruses carrying particular convergent mutations, we can propose a “meta-signature” for the most adaptive amino acid states at 35 sites within 501Y lineage genomes (Table S3; Figure 4). In addition to the 28 convergent mutations that displayed frequency increases between 15 March and 01 June 2021, the meta-signature includes deletion mutations at ORF1a/3675-3677, S/69-70, S/144, and S/241-243 (which, while displaying convergence between the different 501Y lineages, were not amenable to selection analyses) and the convergent signature substitutions L18F, K417N/K and N501Y (which were already at high frequencies in multiple 501Y lineages by 15 March 2021).

Before March 2021 most V1, V2 and V3 viruses respectively carried 10, 13 and 11 of the mutations within this meta-signature. By 01 June 2021 17 different V1 variants (represented by 53 sequenced genomes) matched 13 of the meta-signature sites, two different V2 variants (represented by 20 sequenced genomes) matched 16 of the sites and one V3 variant (represented by 4 sequences) matched 14 of the sites: i.e. in all three lineages viruses were present that had taken three additional mutational steps that converged on the meta-signature.

Given that 19/35 of the meta-signature sites are in the S-gene, these ongoing convergence patterns can be best illustrated by examining convergence on the meta-signature spike S-gene sites within the different 501Y lineages since October 2020 (Figure 5). Whereas the prototype V1, V2 and V3 genomes respectively carried only 4, 4 and 6 of the 19 S-gene mutations in the meta-signature, by 01 June 2021 V1, V2 and V3 variants had arisen that each carried seven of these mutations (Figure 5).

**Figure 5.**
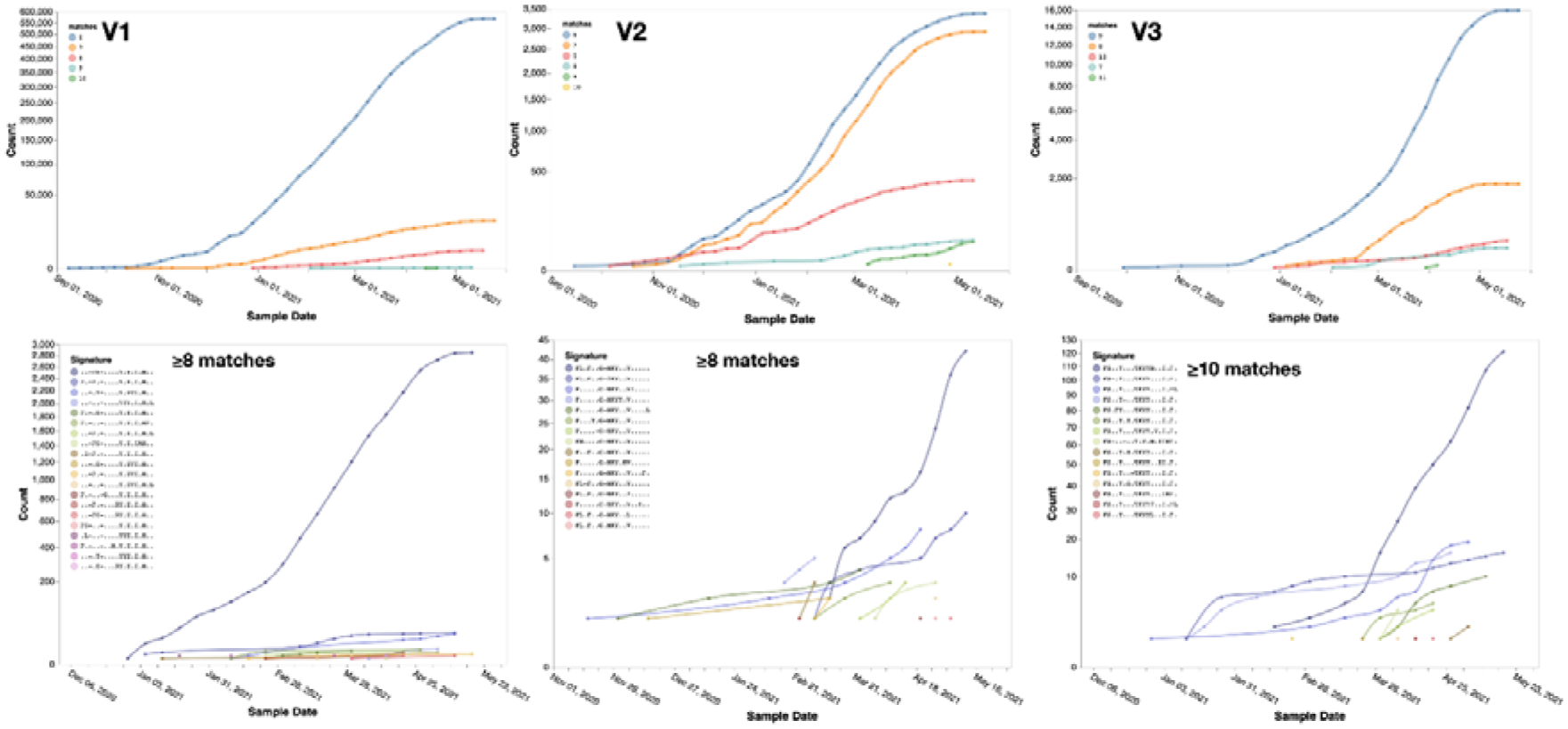
Weekly changes in the counts of sequences displaying multiple convergence mutations at the 19 sites in Spike predicted by our analyses to provide 501Y lineage viruses with selective advantages. This 501Y lineage meta-signature includes the following mutations: 18F, 26R/L/S, 69-70Del, 98F, 138HY, 144Del, 215G/H/V/Y, 241-243Del, 417N/T, 484K, 501Y, 655Y, 681L/R/H, 701V, 716I, 1027I, 1118H, 1176F, 1264L. The matches plots (top row) indicate the numbers of sequenced V1, V2 and V3 genomes carrying a given number of matching mutations at sites on this list: archetypical V1, V2 and V3 sequences respectively have Spike sequences with four, four and six matches. The signature sequence plots (bottom row) indicate the counts of particular V1, V2 and V3 spike sequence haplotypes with the highest numbers of matches and indicate the subsets of mutations in these haplotype sequences. The signature lists included together with these plots indicate the subset of mutations at the 19 convergence list sites that are present in the different Spike haplotype sequences represented in the plots. “.” symbols indicate the absence of a convergence list mutation, “-” symbols indicate the occurrence of convergence list deletion mutations and letters indicate the presence of convergence list amino acid substitutions.

Beyond the three 501Y lineages, we compared degrees of convergence on the 501Y meta-signature of all SARS-CoV-2 genome sequences in GISAID on 01 June 2021 that were classified as belonging to lineages designated by either Public Health England or the US CDC as variants of concern (VOC), variants of interest (VOI) or variants under investigation (VUI; Table S3). All of these lineages had viruses assigned to them that matched at least one of the meta-signature mutations. The lineages containing sequences with the most matches were B.1.526 and B.1.621 (both with modal matches = 5 and maximum = 7), suggesting that they too are possibly scaling the same fitness peak as the 501Y lineage viruses.

Other prominent VOC, VOI and VUI lineages, however, had very few matches. For example the best matched sequences within the B.1.617.2, P.2 and R.1 lineages each had only three matches to the meta-signature (with modal matches in each being one, two, and one, respectively), implying that viruses in these lineages are likely scaling a different fitness peak to the one that the 501Y lineage viruses are on. Curiously, although the modal number of matches to the meta-signature of viruses in B.1.617.1, the sister lineage of B.1.617.2, is only one, this lineage contains some sequences that match the meta-signature at six sites, suggesting that at least some sub-lineages within B.617 are possibly also climbing the same fitness peak as the 501Y lineage viruses.

The non-501Y lineage SARS-CoV-2 isolates that most closely match the meta-signature are found within B.1.620; a lineage first detected in Lithuania (but likely originating in Central Africa) that is presently not considered a VOI, VOC or VUI (Dudas et al., 2021). Whereas the modal number of meta-signature matches for members of B.1.620 is eight (3675-3677Del in ORF1a; 26S, 69-70Del, 241-243Del, 484K, 681H, 1027I, 1118H in the S-gene), some sequences within the lineage have ten matches, suggesting both that the members of this lineage are on the same fitness peak as the 501Y lineage viruses, and that they too are discovering predictable paths to its summit.

We therefore anticipate that the culmination of the currently ongoing evolutionary convergence of 501Y lineage viruses will yield a succession of variants possessing increasing subsets of 501Y lineage meta-signature mutations. The most important issue is not whether we correctly predict the emergence of a super-variant carrying mutations at every one of the 35 meta-signature sites. It is rather that the convergent mutations that are continuing to arise, both in members of the 501Y lineages and those of lineages such as B.1.620, B1.621 and B1.526, imply that all these viruses are presently on, and are actively scaling, the same broad peak in the fitness landscape. Whatever SARS-CoV-2 variants eventually summit that peak could be a considerably bigger problem for us than any we currently know, in that they might have any combinations of increased transmissibility, altered virulence and/or increased capacity to escape population immunity.

Although only time can test the accuracy of this prediction, it should also be possible using *in vitro* evolution to infer some amino acid sequence features at the adaptive summit of this fitness peak. An obvious, albeit potentially controversial, approach would be to use replicated, laboratory infections of either synthesised or sampled live viruses carrying complements of mutations that are representative of the current standing diversity within the V1, V2 and V3 lineages. In the presence of mixed sera from multiple previously infected and/or vaccinated individuals these infections would create the appropriate conditions both for genetic recombination to occur, and for selection to rapidly sort multiple recombination-generated combinations of input immune evasion, cell entry and replication impacting mutations. Although the chimaeras that ultimately dominate these *in vitro* infections will doubtlessly be cell-culture optimized (as opposed to transmission between, and replication within, humans optimized) they should nevertheless carry many combinations of mutations that will be relevant to the continuing pandemic and which should include some of the most concerning mutation combinations that might arise before the pandemic concludes: perhaps particularly so when 501Y lineage viruses start frequently recombining with each other. Even if the concerning combinations that are discovered are only triplets or quartets of mutations, these would still be invaluable hints at what we should start looking for when it comes to trawling the rapidly growing pool of SARS-CoV-2 genomic surveillance data for potential vaccine escape mutants and other potentially problematic variants.

## Methods

### Global SARS-CoV-2 sequence datasets

Since genes/peptides are the targets of selection, unless specified otherwise here and hereafter, all analyses were performed on single genes (e.g. S) or peptide encoding gene segments (e.g. nsp3). We developed an open-source bioinformatics workflow to handle large volumes of sequencing data (>1,000,000 sequences) in a systematic and scalable manner (covid19.galaxyproject.org). Until SARS-CoV-2 emerged in 2019, analyses of natural selection using a few thousand viral sequences would be considered “large scale” (Murrell et al., 2013). Our approach represents a substantial technical advance over the previous state of the art. Because we were also interested in temporal trends in selective pressures, we partitioned all the sequences into three-month intervals based on the date of sampling, and analyzing sliding temporal windows, starting on the 1st of each month from March 2020 to May 2021:

1. We downloaded and curated GISAID sequence data, removed sequences that contained too many ambiguous or unresolved nucleotides, and identified all unique haplotypes for each of the 23 genomic regions that were then analysed individually (*3C, E, endornase, exonuclease, helicase, leader, M, methyltransferase, N, nsp2, nsp3, nsp4, nsp6, nsp7, nsp8, nsp9, nsp10, ORF3a, ORF6, ORF7a, ORF8, RdRp, S*).
2. Each unique haplotype was translated into an amino-acid sequence via a procedure that allows correction of out-of-frame sequencing errors (while not common, there are several thousand sequences in GISAID which have these errors), following which these translated sequences were mapped to the NCBI SARS-CoV-2 genome reference using the *bealign* tool from the BioExt package (github.com/veg/bioext) using the scoring matrix developed for rapidly evolving RNA viruses (Nickle et al., 2007). We did not keep track of insertions relative to the reference genome (this is common practice in the field, since there are no widely circulating strains with evidence of insertions). Therefore, mapping to the reference sequences of individual gene and gene segments generated multiple sequence alignments that were suitable for downstream analyses. These data were directly used for tabulating mutation frequencies and tracking haplotypes with mutations that matched specific mutation signatures (see https://observablehq.com/@spond/spike-trends)
3. Direct comparative analyses of tens or hundreds of thousands of haplotypes is technically very challenging, but is not necessary with respect to extracting the majority of the available selection signal. This is due to two factors. First, much of the variation in individual genomes is either artifactual (sequencing or assembly errors), or not biologically informative (occurs only in a few strains). Second, analyses in such settings need to account for a well-known feature of viral evolution (Poon et al., 2007) where terminal branches include “dead-end” mutation events within individual hosts which, although maladaptive or deleterious at the population level (Pybus et al., 2007), have not been “seen” by natural selection. Mutations that map to internal tree branches on the other hand are far less likely to be severely maladaptive since they must include at least one transmission event. We therefore implemented three layers of compression to reduce the numbers of sequence haplotypes that needed to be subjected to comparative analyses.

1. We did not retain copies of identical sequences. Instead, all identical sequences were represented by a single haplotype. This was because comparative phylogenetic analyses of evolutionary rates do not gain information from the inclusion of identical sequences.
2. We filtered putative sequencing errors and “problematic” sequences. A mutation that occurs in X out of N total sequences (counting all sequences, not just the unique ones) was considered to be an “error” if the binomial probability of observing X or more error mutations at a site was sufficiently high (in our case p > 0.999) assuming a sequencing error rate of 1:10,000. For example, if N = 500,000, then X would be 29. This means that unless a mutation occurred in 30/500,000 or more individual sequences, it would have been treated as an error and replaced with a phylogenetically uninformative gap character (i.e. “-”). One exception to this rule occured when two rare mutations **a** and **b** that would have been filtered out if considered in isolation, occurred together in more than one sequence (i.e. they represented rare linked mutations); we retained such mutations because the probability of coincidental doublets occurring by chance is quadratically small. Additionally, assuming that the distribution of differences from reference sequences across all sequences had a mean, M, and standard deviation, D, we further removed all sequences that had more than M + 5D mutations. These sequences were deemed to be “unusually” mutated and potentially the product of sequencing device contamination, extensive sequencing errors, and/or real/artifactual sequencing assembly-associated recombination.
3. We grouped the remaining filtered haplotypes into clusters based on complete linkage using TN93 distances for computing pairwise sequence similarities (Rhee et al., 2019); sequences were placed in a cluster if and only if all of the pairwise distances between them were ≤d, where **d** is a gene-specific threshold, e.g. d = 0.001 for S. All the ^≤^sequences belonging to the same cluster were represented by a single “median” sequence from the cluster.
4. For example, consider Spike sequences sampled between 01 March 2021 and 31 May 2021. 534,345 sequences passed the initial step 1 filter. Following the step 3a filter, these were reduced to 63,559 unique haplotypes. Following error correction and haplotype recompression in step 3b (where ‘-’ characters were introduced to reflect corrected putative sequencing errors and where ‘-’ matched any resolved characters) 41,103 haplotypes remained. In step 3c these were then compressed down to 5,147 clusters, each yielding a single representative haplotype sequence that was used for downstream selection analyses.
4. We next reconstructed phylogenetic trees for the remaining haplotype sequences using RapidNJ (Simonsen et al., 2008).
5. We used HyPhy v2.5.31 (http://www.hyphy.org/) (Kosakovsky Pond et al., 2020) to perform a series of selection analyses. This version of HyPhy includes many optimizations that were introduced specifically to deal with large SARS-CoV-2 datasets; since March 2020, targeted optimizations for trees with ≥1,000 leaves allowed HyPhy to process 10-25 times as many sequences as earlier versions. We performed SLAC (for substitution mapping (Kosakovsky Pond and Frost, 2005)), FEL (for pervasive positive diversifying and negative selection detection (Kosakovsky Pond and Frost, 2005)), and MEME (for episodic positive diversifying selection detection (Murrell et al., 2015)) analyses that were restricted to considering only mutations mapping to internal branches of inferred trees. These analyses reported p-values and inferred dS and dN rates and ratios for individual codon sites.

### Lineage-specific datasets

We used the RASCL tool (Faria et al., 2021; Lucaci et al., 2021; Tegally et al., 2021) to perform a more detailed analysis of downsampled V1, V2 and V3 gene and gene-segment datasets. For a given gene or gene segment we aligned all sequences from an individual lineage (i.e. V1, V2 or V3) and reference sequences (GISAID unique haplotypes in the corresponding gene/peptide encoding gene segment; Figure 6) to the GenBank reference genome protein sequence for that gene/peptide encoding gene segment using *bealign* with the HIV-BETWEEN-F scoring matrix which is optimized for low-diversity viral sequences.

**Figure 6.**
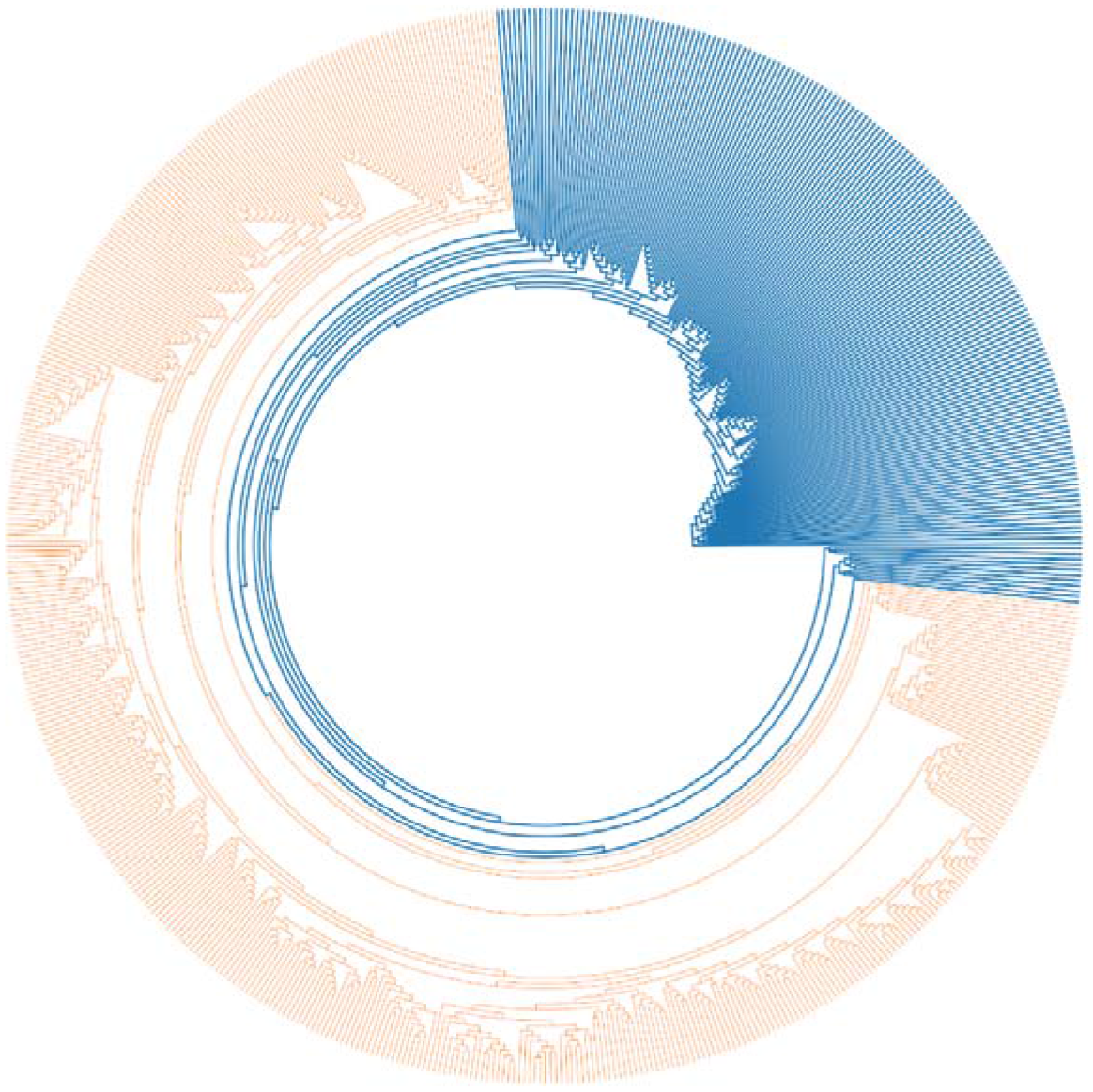
An example of how phylogenetic trees were partitioned into two non-overlapping sets of branches during selection analyses. A foreground clade (here illustrated in orange) is nested within a background tree (illustrated in blue). In our study the foreground clade comprised the subtree relating sequences in either the V1, V2 or V3 lineages to one another and the background clade the tree relating the 501Y lineage sequences to a set of algorithmically selected SARS-CoV-2 reference sequences that were representative of SARS-CoV-2 genetic diversity sampled before October 15th 2020.

Because the codon-based selection analyses that we performed gain no power from including identical sequences, and minimal power from including sequences that are essentially identical, we filtered the V1, V2, V3, and reference (GISAID) sequences using pairwise genetic distances complete linkage clustering with the *tn93-cluster* tool (https://github.com/veg/tn93). All groups of sequences that were within **D** genetic distance (determined using a Tamura-Nei 93 nucleotide substitution model) of every other sequence in the group were represented by a single randomly chosen sequence in the group. We set **D** at *0.0001* for lineage-specific sequence sets, and at *0.0015* for GISAID reference (or “background”) sequence sets (Figure 6). We restricted the reference sequence set to sequences sampled before Oct 15th, 2020 since we were specifically interested in, on a lineage-by-lineage basis, disentangling the impacts of selective processes operating before this date from those operating thereafter. This date approximately marks what appears to have been a major shift in the selective environment within which SARS-CoV-2 is evolving.

We inferred a maximum likelihood tree from the combined sequence dataset with *raxml-ng* using default settings (GTR+G nucleotide substitution model and 20 starting trees). We partitioned internal branches in the resulting tree into two non-overlapping sets used for testing (e.g., orange and blue branches in Figure 6) via encoded annotations made to the Newick tree. Because of low phylogenetic resolution in some of the genes/peptide encoding segments, not all analyses were possible for all segments/genes. In particular this is true when lineage V1, V2 or V3 sequences were not monophyletic in a specific gene/segment, and no internal branches could be labeled as belonging to the foreground lineage.

### Lineage-specific selection analyses

We used HyPhy v2.5.31 (http://www.hyphy.org/) (Kosakovsky Pond et al., 2020) to perform a series of selection analyses. As with the analysis of global datasets we considered only internal-branch mutations in the lineage-specific selection analyses

We performed codon-site-level tests for episodic diversifying (MEME) (Murrell et al., 2015) and pervasive positive or negative selection (FEL) (Kosakovsky Pond and Frost, 2005) on the internal branches of the V1,V2 or V3 clade datasets containing sequences deposited in GISAID by 20 April 2021, to infer the selective dynamics at individual codon-sites across the different SARS-CoV2 genes. Analyses were run with default settings using `--branches Internal` command line flags to restrict dN/dS testing to internal branches only. We only considered as significant those selection signals detected at codon-sites that did not contain nucleotides expressed in multiple frames.

We performed model-based predictions of codons expected to arise is SARS-CoV-2 based on the evolution of related coronaviruses in other host species was determined using the PRIME method http://hyphy.org/w/index.php/PRIME with default settings.

We combined the results of all these analyses using a Python script and visualized them using several open source libraries in ObservableHQ (https://observablehq.com/@spond/n501y-clades).

### Analysis limitations

Selection analyses employed here are not well suited to detecting certain types of selection (e.g. directional selection), for which other specialized techniques can be used (e.g. the DEPS method). Despite our stringent filtering, some of the selection signals detected may have been false positives attributable to sequencing errors, undetected genetic recombination and/or inaccurate phylogenetic inference. Similarly, given the relatively low divergence of SARS-COV-2 genomes, lack of power to detect positive selection (i.e. enough synonymous and non-synonymous substitutions occurring at individual codon sites) was, and will remain, a persistent issue with detecting positive selection in SARS-CoV-2 sequences. Further, all of our comparative analyses were subject to temporal and spatial sampling biases, and country-to-country heterogeneity in time-lags between when viruses were sampled and their sequences became publicly accessible.

### Identification of potentially adaptive convergent mutations

We produced a list of 501Y lineage signature mutation sites at which mutations arising in viruses of any one of the 501Y lineages during or before March 2021 converged on the signature mutation states of viruses in another 501Y lineage. To this list of sites we added a list of non-signature mutation sites at which convergent mutations occurring before March 2021 were observed between two or more different 501Y lineages that incurred enough convergent non-synonymous mutations along internal tree branches (i.e. excluding mutations mapping to terminal tree branches) to trigger positive selection signals with associated MEME or IFEL p-values <0.05. We then tested this combined “convergence list” for evidence of its constituent convergent mutations having more than doubled in frequency within V1, V2 or V3 sequences sampled between 15 March and 01 June 2021 relative to frequencies seen within these lineages before that time. We then repeated this test with 501Y lineage signature mutation sites at which convergent mutations had not been detected prior to March 2021. Finally, all convergent mutations analysed in these tests that (1) more than doubled in frequency within individual 501Y lineages between 15 March 2021 and 01 June 2021, and (2) occurred at signature/non-signature mutation sites displaying significant signals of positive selection in either the global SARS-CoV-2 sequence dataset (IFEL p-value < 0.05) or any one of the lineage-specific datasets (MEME/IFEL p-values < 0.05) were identified as the mutations most likely to positively impact the fitness of the 501Y lineage viruses. This list of mutations was merged with the list of deletion mutations that characterize the different 501Y lineages and the three cardinal 501Y lineage signature mutations, L18F, K417N/K and N501Y which, due to already high frequencies in multiple 501Y lineages could not have doubled in frequency between 15 March and 01 June 2021. This final mutation list is what we called the 501Y meta-signature.

## Data Availability

All Data used was obtained from GISAID (which is credited in the manuscript)

## Acknowledgements

We gratefully acknowledge all of the authors from the originating laboratories responsible for obtaining the specimens and the submitting laboratories where genetic sequence data were generated and shared via the GISAID Initiative, on which this research is based (Table S2).

DPM is funded by the Wellcome Trust (222574/Z/21/Z)

SLKP was supported by grants from the U.S. National Institutes of Health [R01 AI134384 (NIH/NIAID), R01 AI140970 (NIH/NIAID)], and a RAPID award from the US National Science Foundation 2027196 (NSF/DBI,BIO).

DLR is funded by the Medical Research Council (MC_UU_1201412) and Wellcome Trust (220977/Z/20/Z).

OAM is funded by the Wellcome Trust (206369/Z/17/Z).

COG-UK is supported by funding from the Medical Research Council (MRC) part of UK Research & Innovation (UKRI), the National Institute of Health Research (NIHR) and Genome Research Limited, operating as the Wellcome Sanger Institute.

PL acknowledges funding from the European Research Council under the European Union’s Horizon 2020 research and innovation programme (grant agreement no. 725422-ReservoirDOCS), the EU grant 874850 MOOD and the Wellcome Trust through project 206298/Z/17/Z.

JOW was supported by an NIH-NIAID R01 AI135992.

SEJ and HT are supported by H3ABioNet, an initiative of the Human Health and Heredity in Africa Consortium (H3Africa) funded by the National Human Genome Research Institute of the National Institutes of Health under Award Number U24HG006941.

The Network for Genomic Surveillance South Africa (NGS-SA) is supported by the Strategic Health Innovation Partnerships Unit of the South African Medical Research Council, with funds received from the South African Department of Science and Innovation.

GWH is supported by a grant from the US National Institutes of Health (1U01Al152151-01)

## Supplementary Figures

**Figure S1.**
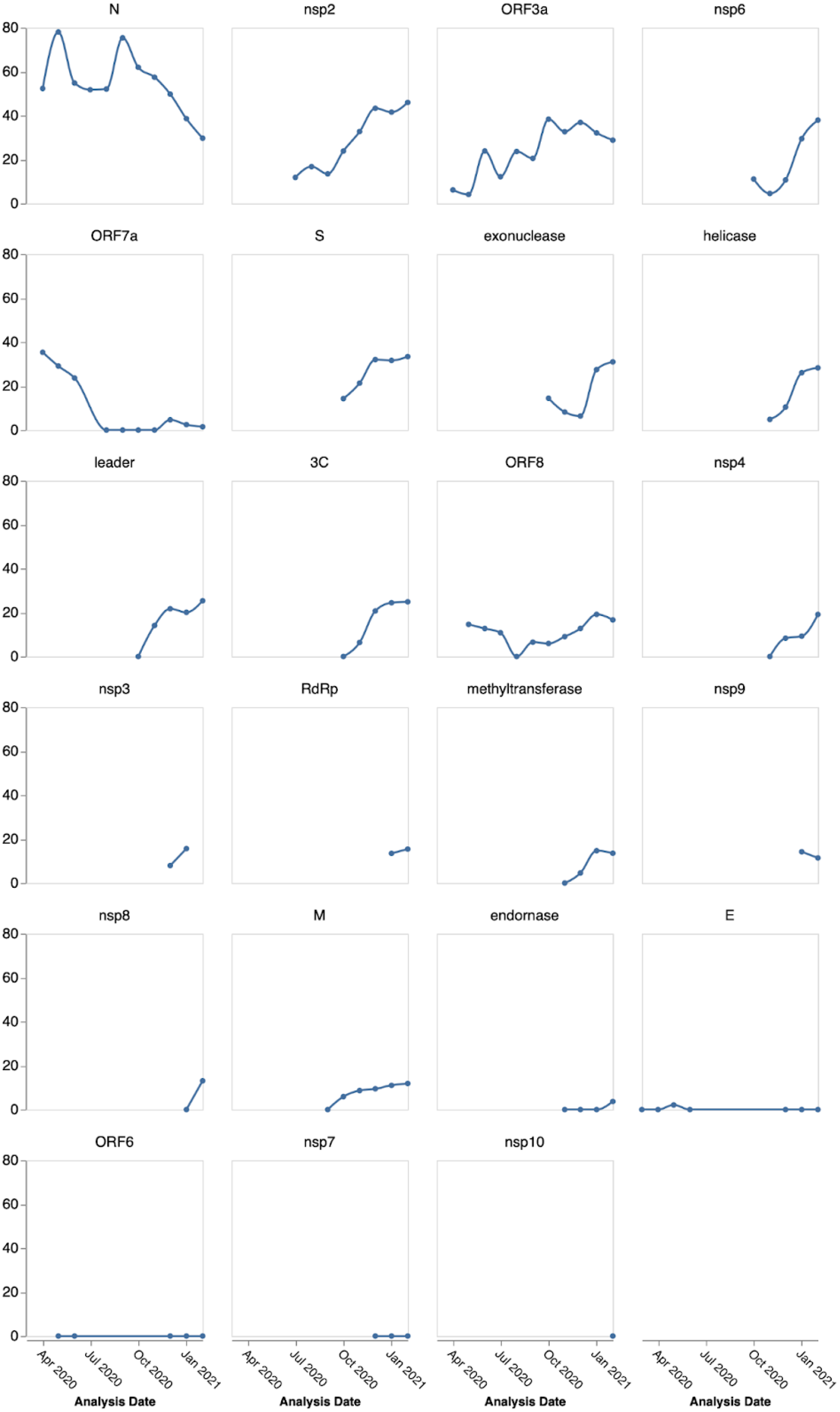
Signals of positive selection at individual codon-sites that were detectable with the FEL method at different times between March 2020 and February 2021, applied to sequences sampled over 90-day intervals; the plotted date shows the end of the 90-day period. Genes with associated trees that have a total length shorter than 0.5 subs/site for a given time-period are not shown. Note that for the ORF3a and the N gene the interpretation of positive selection signals is complicated by the fact that each of these genes encompasses multiple smaller genes that are expressed in different reading frames. This is because synonymous substitutions in the ORF3a and N reading frames will be non-synonymous substitutions in the reading frames of the smaller genes that they encompass (and *vice versa*): a situation that is expected to inflate positive selection signals.

**Figure S2.**
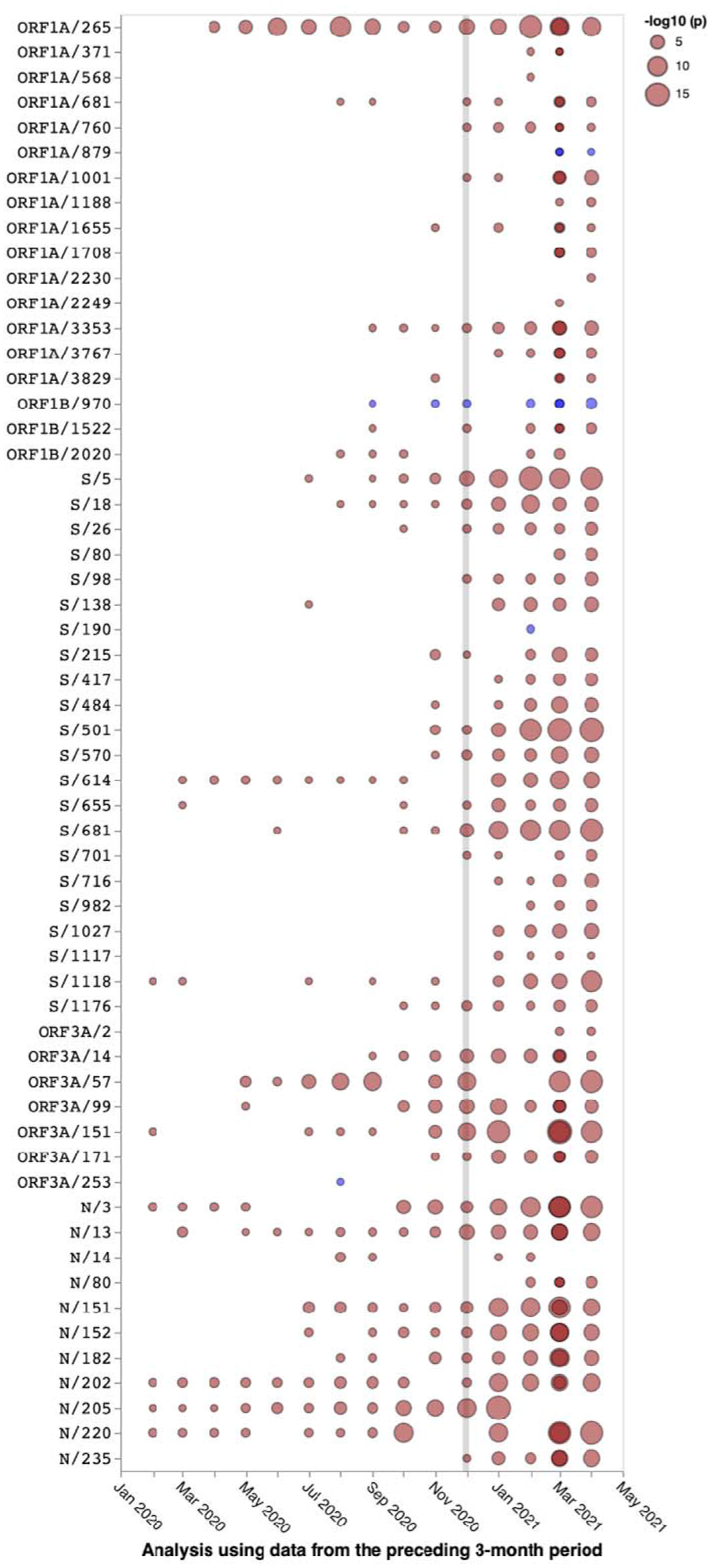
Global selection trends at sites in 501Y lineage viruses that are either signature mutations or which, on 20 April 2021, displayed evidence of both lineage-specific positive selection on internal tree branches and mutational convergence between viruses in different 501Y lineages. Red dots indicate positive selection and blue dots indicate negative selection in the global SARS-CoV-2 dataset. The sizes of the dots indicate the strength of the positive/negative selection signals. Selection signals indicate those detected when considering only the sequences sampled in the preceding three months.

## Supplementary Tables

**Table S1.**
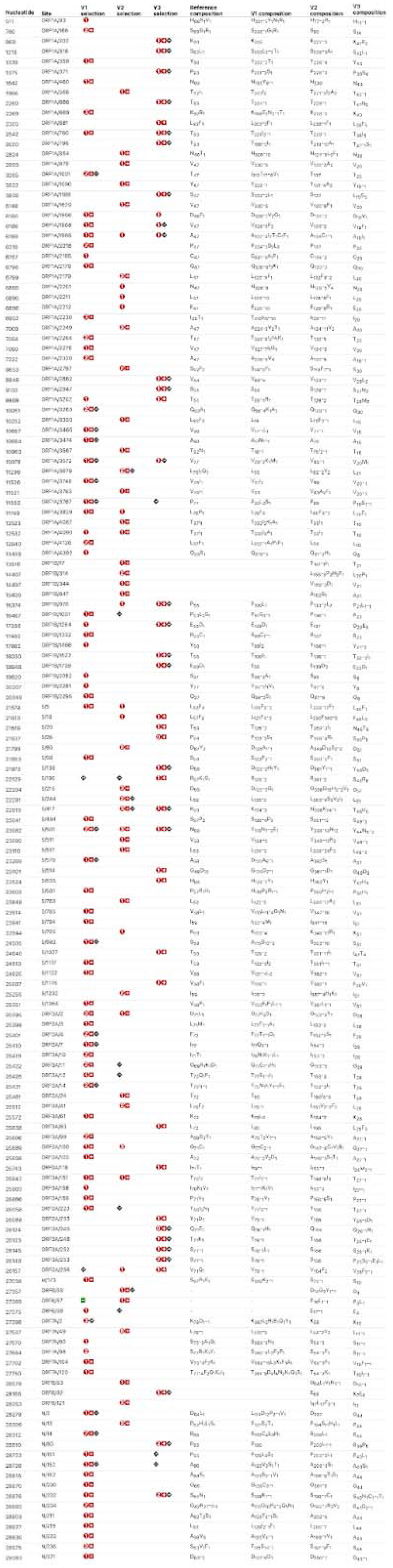
Sites evolving under positive selection in 501Y lineage-specific datasets. Also indicated are the alternative encoded amino acids at these sites with subscripts associated with the encoded amino acid states indicating numbers of analysed sequences encoding those amino acids.

**Table S2.** Changes in mutation frequencies at signature and/or convergent mutation sites within 501Y lineage viruses between 15 March and 01 June 2021 that are detectably evolving under positive selection in individual lineages and/or in the global SARS-CoV-2 dataset

| ORF(Gene) | Inter-lineage convergence detected in April and **June** | Fold-increase in lineage V1 | Fold-increase in lineage V2 | Fold-increase in lineage V3 |
| --- | --- | --- | --- | --- |
| ORF1a/265I(nsp2/85I) | V1,V2 | 2.2 | Fixed |  |
| ORF1a/681F(nsp2/501F) | V1,V2,V3 | 1.6 | 3.0 | <0.1 |
| ORF1a/1001I(nsp3/183I) | **V1,V2** | Fixed | 2.2 |  |
| ORF1a/1188L(nsp3/370L) | V1,V3 | 2.5 |  | Fixed |
| ORF1a/1655N(nsp3/837N) | **V1,V2,V3** | 2.5 | Fixed | 6.6 |
| ORF1a/1708D(nsp3/890D) | **V1,V2,V3** | Fixed | 15.8 | >12.0 |
| ORF1a/3829F(nsp6/260F) | V1,V2,V3 | 2.2 | 0.5 | 1.4 |
| ORF1b/970L(Helicase/47L) | V1,V2,V3 | 1.7 | 0.2 | <0.1 |
| ORF1b/970S(Helicase/47S) |  | 2.2 |  |  |
| ORF1b/1522I(Helicase/599I) | V1,**V2**,V3 | 1.9 | 12.7 | 4.8 |
| ORF1b/2020Y(Exonuc/496Y) | V1,**V2**,V3 | 0.7 | 2.2 | <0.1 |
| S/26S | V1,**V2**,V3 | 0.8 | >13.0 | Fixed |
| S/26L |  |  | 11.1 |  |
| S/98F | V1,V2,**V3** | 2.5 | 5.3 | >6.0 |
| S/138Y | V1,V2,V3 | 2.1 | 0.8 | Fixed |
| S/215G | V1,V2 | <0.1 | Fixed |  |
| S/215Y |  | 2.1 |  |  |
| S/215C |  |  | >6.4 |  |
| S/484K | V1,V2,V3 | 3.7 | Fixed | Fixed |
| S/655Y | V1,V2,V3 | 2.3 | 0.2 | Fixed |
| S/681H | V1,V2,V3 | Fixed | 0.6 | 21.0 |
| S/681R |  | 2.2 |  |  |
| S/701V | V1,V2 | 2.4 | Fixed |  |
| S/701S |  | 3.1 |  |  |
| S/716I | V1,V2,**V3** | Fixed | 3.7 | >13.5 |
| S/1027I | V2,V3 |  | >44.0 | Fixed |
| S/1118H | **V1,V2,V3** | Fixed | 4.0 | >21.0 |
| S/1176F | **V2,V3** |  | 19.5 | Fixed |
| S/1264L | V1,V2,**V3** | 2.3 | 0.4 | 1.4 |
| ORF3a/57H | V1,V2,**V3** | 1.4 | Fixed | 2.8 |
| ORF3a/171L | V1,V2,**V3** | 0.9 | Fixed | 11.9 |
| E/71L | **V1,V2** | 1.1 | Fixed |  |
| E/71T |  | 5.8 |  |  |
| E/71R |  | >10 |  |  |
| N/205I | V1,V2,**V3** | 0.8 | Fixed | 1.2 |
| N/205- | **V1,V3** | 1.4 |  | 3.0 |
| N/235F | V1,V2,**V3** | Fixed | 2.3 | >10.0 |
| N/235L |  | 2.9 |  |  |

**Table S3.** Mutations comprising the 501Y lineage meta-signature

| Mutation (gene segment coordinate) | Positive selection G=Global (in lineage VX) | Inter-lineage convergence | Maximum fold increase between March and June 2021 (lineage) |
| --- | --- | --- | --- |
| ORF1a/265I(nsp2/85I) | G | V1,V2 | 2.2 (V1) |
| ORF1a/681F(nsp2/501F) | G (V3) | V1,V2,V3 | 3.0 (V2) |
| ORF1a/1001I(nsp3/183I) | G (V1) | V1,V2 | 2.2 (V2) |
| ORF1a/1188L(nsp3/370L) | G (V3) | V1,V3 | 2.5 (V1) |
| ORF1a/1655N(nsp3/837N) | G | V1,V2,V3 | 6.6 (V3) |
| ORF1a/1708D(nsp3/890D) | G | V1,V2,V3 | 15.8 (V2) |
| ORF1a/3676-(nsp6/107-) | G | V1,V2,V3 | -- |
| ORF1a/3829F(nsp6/260F) | G (V1) | V1,V2,V3 | 2.2 (V1) |
| ORF1b/970LS(Helicase/47LS) | (V3) | V1,V2,V3 | 2.2 (V1) |
| ORF1b/1522I(Helicase/599I) | G (V3) | V1,V2,V3 | 12.7(V2) |
| ORF1b/2020Y(Exonuc/496Y) | (V3) | V1,V2,V3 | 2.2 (V2) |
| S/26LRS | G (V3) | V1,V2,V3 | >13.0 (V2) |
| S/69- | G | V1,V2,V3 | 1.7 (V2) |
| S/98F | G (V1) | V1,V2,V3 | >6.0 (V3) |
| S/138HY | G (V3) | V1,V2,V3 | 2.1 (V1) |
| S/144- | G | V1,V2,V3 | 2.3 (V2) |
| S/215GHVY | G (V2) | V1,V2 | 2.1 (V1) |
| S/243- | G | V1,V2,V3 | <0.1 (V1,V3) |
| S/417NT | G (V2,V3) | V2,V3 | <0.1 (V1) |
| S/484K | G | V1,V2,V3 | 3.7 (V1) |
| S/501Y | G (V1,V2,V3) | V1,V2,V3 | -- |
| S/655Y | G (V3) | V1,V2,V3 | 2.3 (V1) |
| S/681LRH | G (V1) | V1,V2,V3 | 21.0 (V3) |
| S/701V | G | V1,V2 | 2.4 (V1) |
| S/716I | G | V1,V2,V3 | >13.5 (V3) |
| S/1027I | G (V3) | V2,V3 | >44.0 (V2) |
| S/1118H | G | V1,V2,V3 | >21.0 (V3) |
| S/1176F | G | V2,V3 | 19.5 (V2) |
| S/1264L | (V1) | V1,V2,V3 | 2.3 (V1) |
| ORF3a/57HLY | G | V1,V2,V3 | 2.8 (V3) |
| ORF3a/171L | G | V1,V2,V3 | 11.9 (V3) |
| E/71LRT |  | V1,V2 | >10.0 (V1) |
| N/205I- | G | V1,V2,V3 | 3.0 (V1) |
| N/235FL | G (V1) | V1,V2,V3 | >10.0 (V3) |

**Table S4.** Sequences within different SARS-COV-2 lineages of concern, lineages of interest and lineages under investigation that have most converged on the 501Y lineage meta-signature. Indicated in green are mutations that converge on the 501Y lineage meta-signature which are not signature/lineage-defining mutations in the lineages from which the sequences were drawn. https://docs.google.com/spreadsheets/d/1QT7NMDYa8VzmEAkrskeKk3ruE5SIU7CxZS3j1fjCSio/edit?usp=sharing

